# Dynamics of non-household contacts during the COVID-19 pandemic in 2020 and 2021 in the Netherlands

**DOI:** 10.1101/2022.10.19.22281248

**Authors:** Jantien A. Backer, Laurens Bogaardt, Philippe Beutels, Pietro Coletti, W. John Edmunds, Amy Gimma, Cheyenne C. E. van Hagen, Niel Hens, Christopher I. Jarvis, Eric R. A. Vos, James Wambua, Denise Wong, Kevin van Zandvoort, Jacco Wallinga

## Abstract

The COVID-19 pandemic was in 2020 and 2021 for a large part mitigated by reducing contacts in the general population. To monitor how these contacts changed over the course of the pandemic in the Netherlands, a longitudinal survey was conducted where participants reported on their at-risk contacts every two weeks, as part of the European CoMix survey. The survey included 1659 participants from April to August 2020 and 2514 participants from December 2020 to September 2021. We categorized the number of unique contacted persons excluding household members, reported per participant per day into six activity levels, defined as 0, 1, 2, 3-4, 5-9 and 10 or more reported contacts. After correcting for age, vaccination status, risk status for severe outcome of infection, and frequency of participation, activity levels increased over time, coinciding with relaxation of COVID-19 control measures.

## Introduction

From early 2020 onwards, the COVID-19 outbreak grew into a global pandemic, taking only a few months to affect nearly all countries worldwide. SARS-CoV-2 virus that causes COVID-19 is transmitted between persons during contact events with physical proximity, i.e. at-risk contacts. Reducing transmission with non-pharmaceutical measures can be achieved by lowering the transmission probability per contact, e.g. by maintaining a safe distance from each other or using protective equipment such as a face mask, and by reducing the number of contacts per person, e.g. by closing schools, theaters and restaurants, teleworking or inviting fewer people at home.

In the Netherlands, the first nation-wide lockdown was imposed on March 15th, 2020 involving all of the above measures. It was lifted stepwise on May 11th (partly opening of schools), June 2nd (further opening of schools, museums and terraces open a.o.) and July 6th (bars and restaurants open a.o.). The rise of the Alpha variant in winter 2020/2021 called for an even stricter lockdown including an evening curfew. The measures were relaxed from April 28th, 2021 onwards as vaccination coverages were increasing. It is not known how the change in control measures affected the number of at-risk contacts in the Dutch general population.

To learn about the effect of control measures on contact behaviour, many countries set up contact surveys^1–9^. The POLYMOD study^10^ served as a blueprint for these as it did for many pre-COVID-19 studies^11^. But while these pre-COVID-19 studies aimed to measure ‘normal’ contact behaviour, the unprecedented measures that were taken during the pandemic gave a unique opportunity to determine contact behaviour under restricted conditions. Understanding contact patterns between age groups is especially important when restrictions affect age groups differently. Physical distancing measures were often aimed at reducing contacts with old or frail persons, as they have a higher chance of hospitalisation and death after infection^12–14^. During the first wave of the COVID-19 pandemic, the in-person contacts of elderly persons decreased^15^, and also surveys in the general population showed a substantial reduction of contacts^1–9^.

As part of a European project, the CoMix contact survey was set up in March/April 2020 in Belgium, the UK, and the Netherlands, later followed by 19 other European countries^16^. In this contact study, participants were asked to regularly report their contact behaviour, i.e. the number of unique persons they had contact with in certain locations on the day preceding the survey day, as well as some characteristics of the contacted persons such as age and sex. Participants were asked to report only at-risk contacts, specifically face-to-face conversational contacts (within 1.5 meters) and/or physical contacts. What sets the CoMix survey apart from other contact surveys is that it uses a standardised questionnaire in multiple countries and is repeated at regular intervals in the same participants.

At the same time the CoMix survey started, a large-scale population-based SARS-CoV-2 serosurveillance study was initiated in the Netherlands^17,18^. This Pienter Corona (PiCo) study was repeated every 4 to 6 months in a representative sample of the Dutch population involving a few thousand participants of all ages per survey round, and is still ongoing. It also included a contact question where the participants were asked to report the number of unique persons they contacted the day before in specific age groups. An early analysis of the first two survey rounds in 2020 showed that the first lockdown decreased the number of community contacts per participant on average by 76% compared to pre-COVID behaviour^4^. The PiCo survey rounds are too far apart to closely monitor the contact behaviour over time, but will be used to compare to the CoMix survey results at certain points in time.

Our aim is to describe how control measures affected the contact behaviour of the Dutch general population, using the CoMix data that was collected in the Netherlands during the survey periods in 2020 and 2021. Although contacts with household members are thought to be more intensive than contacts with non-household members, they are not subject to any of the control measures. For this reason, we will focus on the number of contacts per participant outside the household because these can be affected by the control measures. The results are compared to four survey rounds of the PiCo survey that coincided with the CoMix study periods.

## Methods

### CoMix participant recruitment and survey design in the Netherlands

This contact survey was part of the CoMix study that was conducted in several EU countries and the UK^16^. Participants were recruited by the market research company Ipsos-MORI, and filled out the contact survey online. The study was carried out in two survey periods: 8 survey rounds from 16 April to 5 August 2020, referred to as the 2020 series, and 20 survey rounds from 23 December 2020 to 22 September 2021, referred to as the 2021 series. At the start of the 2020 series, 1500 participants of 18 years and older were recruited, reflecting the age distribution of the Dutch population. An overall drop-out rate of 10% per round was allowed, otherwise additional participants were recruited for the following rounds. However, the drop-out rates differed per age group leading to very few participants in the youngest age group of 18-24 in the last rounds of the 2020 series (Fig 1). For this reason, a preferential sampling scheme was adopted at the start of the 2021 series, oversampling the younger age groups. In total 1200 participants of 18 years and older were included and additionally 300 children between 0 and 17 years of age were included by asking adult participants to fill out the contact survey for one of their children. After 10 rounds, the age cohorts were supplemented to include 1500 participants in total again in May 2021.

**Figure 1.**
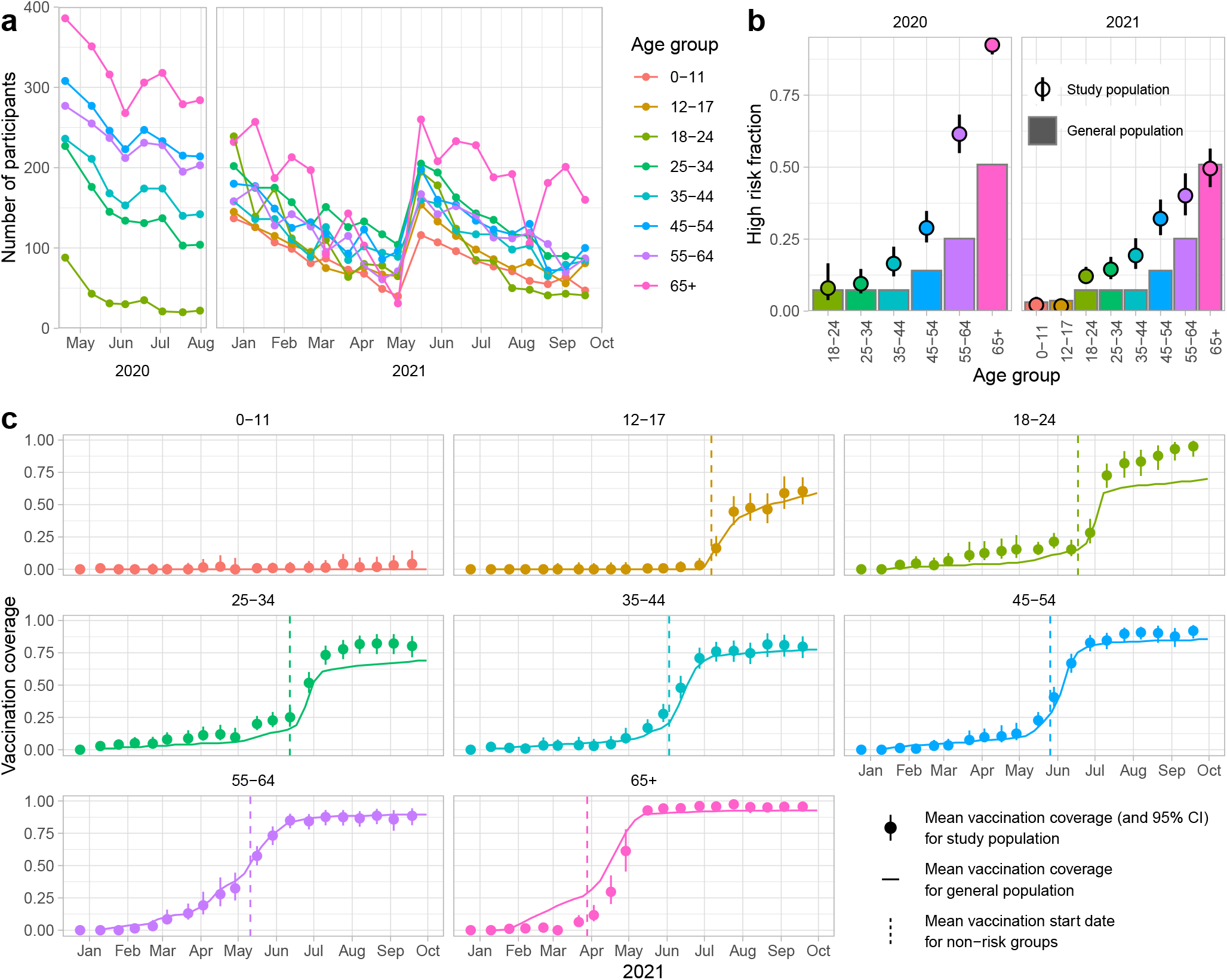
Characteristics of study population, compared to general population. **a** Number of participants included per survey round in the 2020 and 2021 series, by age group. After 10 survey rounds in May 2021, the study population was supplemented to meet the target numbers of the first survey round again. **b** The fraction of high risk participants of the 2020 and 2021 series (points with 95% confidence interval), and of the general population^19^ (bars). Only participants with unambiguous risk status are included. **c** Vaccination coverage in 2021 per age group of the study population (points with 95% confidence interval) and of the general population^20^ (line). The dashed line denotes the average date when non-risk groups were invited for vaccination. The dashed line for the 55-64 age group is in fact the average for 55-59 year olds, as 60-64 year olds were invited by their general practitioner.

The contact survey was repeated every two weeks and a survey round lasted at most 7 days, except for the survey rounds where many new participants were recruited that were allowed more time to fill out the questionnaire. In each survey round participants were asked about characteristics that can change over time, such as risk perception, risk status and - in the 2021 series - vaccination status against COVID-19. High risk participants were people with self-reported chronic respiratory disease, chronic heart disorder, chronic kidney disease, diabetes, reduced resistance to infection (due to illness or medication), morbid obesity (BMI over 40), and pregnant women, i.e. reflecting the medical indication for influenza vaccination. Missing data in the risk status of a participant were completed with the most common risk status reported over the survey rounds by the participant. The risk status of participants could change by survey round. The fraction of high risk participants is calculated per age group using only the participants with unambiguous risk status, and compared to the population fraction with a medical indication for influenza vaccination^19^.

The 2021 series coincided with the roll-out of the COVID-19 vaccination campaign. Health care workers, persons with a high medical risk and care home residents were vaccinated with priority, while the non-risk groups were vaccinated from elderly to young. Most persons were vaccinated with either BionTech/Pfizer, Moderna or to a smaller extent Janssen vaccine by the Municipal Health Services, except for 60-64 year olds who were vaccinated at an early stage with AstraZeneca by their general practitioner. Participants are considered to be vaccinated after they have reported to have had at least one vaccination dose. The fraction of vaccinated participants is compared to the vaccination coverage of one vaccine dose in the general population over time^20^.

In the main part of the survey, participants reported their contact behaviour of the previous day. For each unique person they had conversational or physical contact with between 5am on the previous day and 5am of the survey day, participants filled out the characteristics of the contacted person, such as age group and gender, as well as duration and location of the contact event. Multiple contact events with the same person were to be considered as one contact, and only contact events with a close physical proximity were to be reported, i.e. excluding online or phone contacts. Besides these individual contacts, it was also possible to report group contacts from the third survey round onwards in late May 2020. Group contacts were reported as the total number of contacted persons in broad age groups (0-17, 18-64, 65+) at work, school or another location. More details on the CoMix study and questionnaire can be found in previous publications^16,21,22^.

The Medical Research Ethics Committee (MREC) NedMec confirmed that the Medical Research Involving Human Subjects Act (WMO) does not apply to the CoMix study in the Netherlands (research protocol number 22/917). Therefore an official approval of this study by the MREC NedMec is not required under the WMO. The study was in accordance with relevant GDPR guidelines and regulations. Participants gave informed consent to participate in the study before taking part.

### Analysis of number of contacts

The participants are stratified into three age groups (0-17, 18-64, 65+) and two series (2020, 2021). Only contacts with non-household members are included, as these are most likely to change over the course of the study period. Twelve occasions were excluded where participants reported more than 1000 contacts in a single survey round, and 58 participants were excluded that participated in four or more rounds but did not report any contact. These excluded participants did not have a significantly different age than the included participants, but they did have a significantly smaller household size.

The distribution of the number of contacts per participant per round had a heavy tail, which can result in unstable estimates of the variance^23^. Instead of using a parametric distribution, we define activity levels of 0, 1, 2, 3-4, 5-9, ≥10 contacts per participant per round and analyse these using ordinal regression. The activity levels are used as variables of a generalised additive model with fixed effects for participant vaccination status, participant risk status, weekends and (school) holidays, cubic splines for calendar time, participant age, participant round (i.e. the n^th^ time a participant participated), and a random intercept for each participant. The statistical model can be described as:

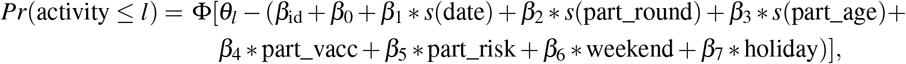

where Φ is the logistic function, *θ*_*l*_ are the thresholds for the activity levels *l, β*_id_ are the random intercepts and the *s*() function indicates that splines are used. The fits are assessed by explained deviance and by comparing the fitted activity levels to the observations.

Next, the fitted results are used in a synthetic population that has the same size as the study population, but reflects the general population, with respect to age^24^, risk status^19^ and vaccination status^20^ at each day of the study periods. Participant id’s are sampled with replacement from the study population, and participant round is set to 1 for each day during the study period. 200 parameter sets are sampled from the estimated model parameters, and combined with 200 synthetic populations, to capture both the uncertainty of the model parameters and the uncertainty caused by the sample size of the study population. The predicted activity levels of the synthetic population are compared to the activity levels measured in PiCo survey rounds 1 (Apr 2020), 2 (Jun 2020), 4 (Feb 2021) and 5 (Jun 2021).

All analyses are performed in R version 4.1.3^25^ using the *mgcv* package^26^ with family *ocat* for ordered categorical variables. Code is available on Github^27^, and data is published on Zenodo^28^ in the standardised format of *socialcontactdata*.*org*.

## Results

### Study population

In total, 1659 participants were included in the 2020 series, and 2514 participants in the 2021 series. Participants were equally distributed over men and women in 2020, while female participants were slightly overrepresented in 2021 (Tab. 1). Household sizes of 2 persons were most common (Tab. 1). Each series started with a target population of 1500 participants, and the number of participants declined in each survey round (Supplementary Tab. S1, Supplementary Fig. S1). In 2020 the drop-out rate was on average 4% per round in the oldest age group and 14% in the youngest age group; in 2021 drop out rates were more comparable for all age groups (Fig. 1A).

**Table 1.** Participants in the CoMix survey in the Netherlands in 2020 and 2021. ^a^ Characteristics for 2020 general population based on 18+ population to match study population. ^b^ Percentage of risk status in study population only calculated for participants with unambiguous risk status to allow comparison with general population. ^c^ Expected household size distribution per Dutch citizen, based on Dutch households consisting of one (38%), two (33%), three (12%), four (12%) and five or more (5%) persons^4^

| 2020 series |  | n | % | reference<sup>a</sup> (%) |
| --- | --- | --- | --- | --- |
| Total |  | 1659 |  |  |
| Age group | 18-24 | 84 | 5 | 11 |
|  | 25-34 | 236 | 14 | 16 |
|  | 35-44 | 257 | 15 | 15 |
|  | 45-54 | 337 | 20 | 18 |
|  | 55-64 | 312 | 19 | 17 |
|  | 65+ | 433 | 26 | 24 |
| Gender | Female | 814 | 49 | 51 |
|  | Male | 845 | 51 | 49 |
| High risk | mixed | 359 | <sup>b</sup> |  |
|  | no | 744 | 58 | 78 |
|  | yes | 540 | 42 | 22 |
|  | missing | 16 | <sup>b</sup> |  |
| Household size | 1 | 409 | 25 | 17<sup>c</sup> |
|  | 2 | 716 | 43 | 31 |
|  | 3 | 241 | 15 | 17 |
|  | 4 | 203 | 12 | 23 |
|  | 5+ | 90 | 5 | 12 |
| 2021 series |  | n | % | reference (%) |
| Total |  | 2514 |  |  |
| Age group | 0-11 | 240 | 10 | 12 |
|  | 12-17 | 290 | 12 | 7 |
|  | 18-24 | 511 | 20 | 9 |
|  | 25-34 | 378 | 15 | 13 |
|  | 35-44 | 254 | 10 | 12 |
|  | 45-54 | 271 | 11 | 14 |
|  | 55-64 | 230 | 9 | 14 |
|  | 65+ | 340 | 14 | 20 |
| Gender | Female | 1383 | 55 | 50 |
|  | Male | 1131 | 45 | 50 |
| High risk | mixed | 385 | <sup>b</sup> |  |
|  | no | 1720 | 81 | 81 |
|  | yes | 400 | 19 | 19 |
|  | missing | 9 | <sup>b</sup> |  |
| Household size | 1 | 573 | 23 | 17<sup>c</sup> |
|  | 2 | 766 | 30 | 31 |
|  | 3 | 444 | 18 | 17 |
|  | 4 | 496 | 20 | 23 |
|  | 5+ | 235 | 9 | 12 |

The fraction of self-reported high risk adult participants was higher than in the general population, especially in the 2020 series (Fig. 1B). The vaccination coverage of the study population increased over time largely in agreement with the general population (Fig. 1C). The participants in the age groups of 18-34 years achieved a higher vaccination coverage at an earlier stage than the general population. The participants in the age group of 65 years and older lagged behind the general population.

### Contact behaviour

The distribution of the number of contacts reveals that the adult and elderly age groups in both series exhibit heavy tails (Fig. 2). The fitted lines on log-log scale (inset in Fig. 2) for these four groups have slopes ranging from -1.1 to -1.5. These values indicate that the estimates of the sample mean are stable but the estimates of the sample variance are not^23^, which makes the choice to model the number of contacts with a parametric distribution less obvious. The number of contacts in the child age group lacks a heavy tail, because of the abundance of children reporting contact numbers in the range of a class size. The numbers of contacts per participant per round are categorized in activity levels of 0, 1, 2, 3-4, 5-9, ≥ 10 contacts per participant per round.

**Figure 2.**
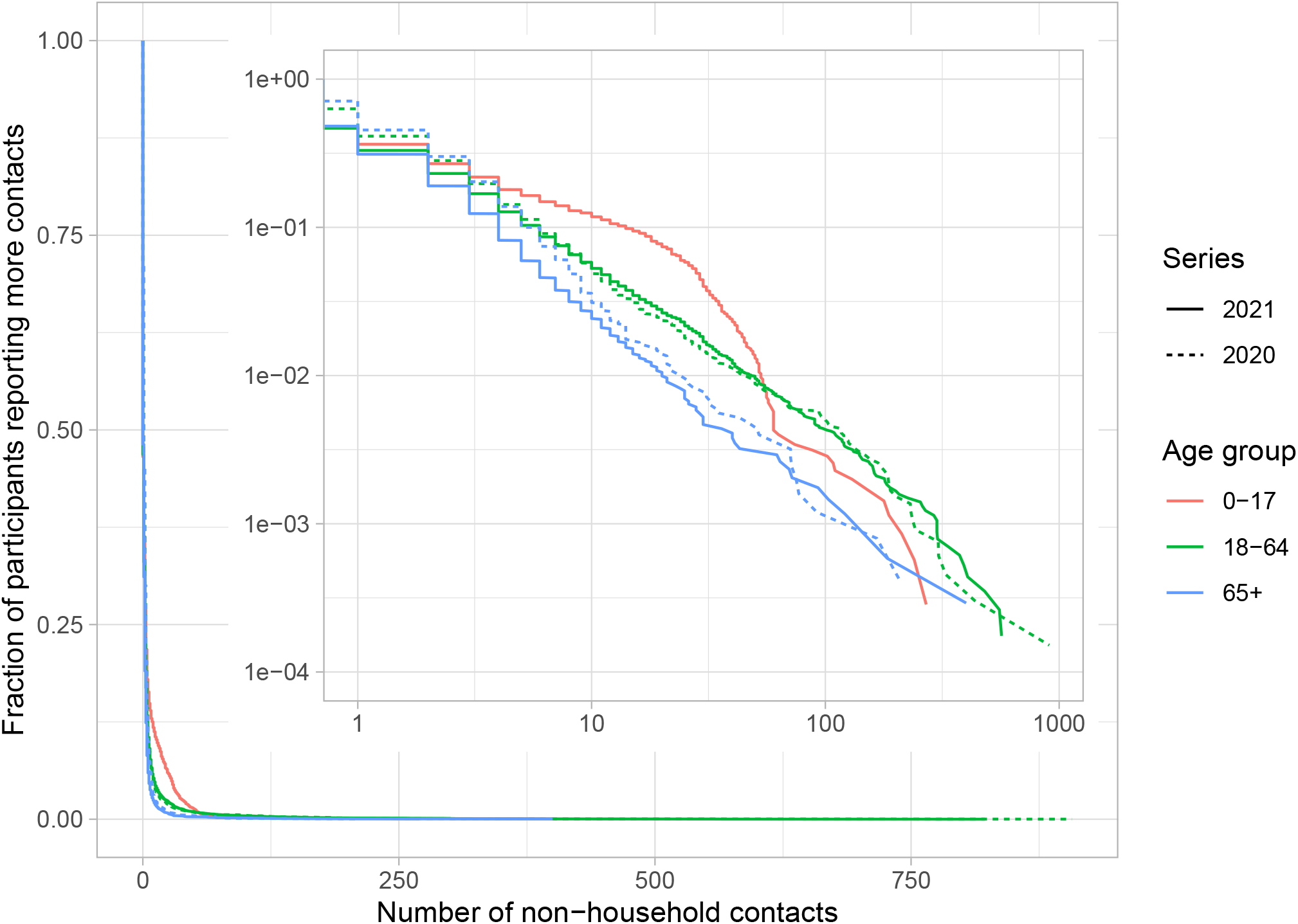
Distribution of number of contacts per participant per round excluding household members. Distribution by series (line type) and age group (color), with complementary cumulative distribution function on y-axis. The inset shows the same plot on log-log scale.

Separate analyses are carried out for the different series and age groups. Of the fixed effects only the weekends significantly reduces the activity levels in all analyses (Supplementary Tab. S2). For the youngest age group, also holidays significantly reduce the activity levels as they also include school holidays. Activity levels increase over time during the two series, indicating that the underlying number of non-household contacts per participant increases (Supplementary Fig. S2), corresponding to the lifting of COVID-19 control measures. In all analyses the activity levels decrease with participant round. This means that participants tend to report fewer contacts the more often they participate. This is especially the case for the first few rounds, after which the effect stabilises.

The deviance explained by this model ranges from 14% to 19% (Supplementary Tab. S2). Using these model results the fitted activity levels in the study population are compared to the data (Fig. 3). With the exception of some rounds, notably the first round in 2020 and the fourth round in 2021, the fitted activity levels agree well with the observed activity levels.

**Figure 3.**
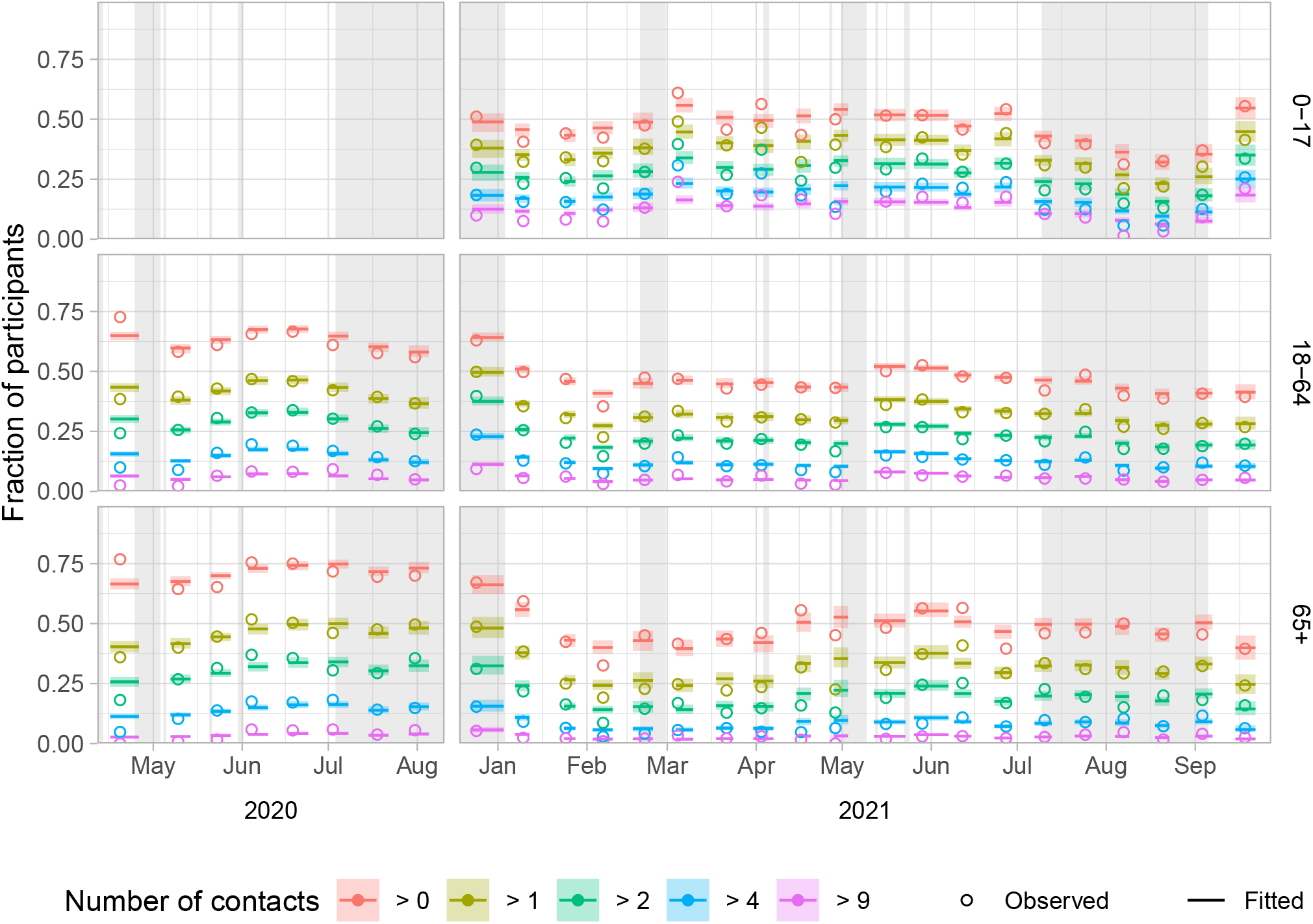
Fitted and observed activity levels over time by series (columns) and age group (rows). Activity levels are shown as the fraction of participants that report more than a certain number of non-household contacts per day. With five limits (>0, >1, >2, >4 and >9) six activity levels are defined, e.g. the fraction between the limits of >2 and >4 is the activity level that represents 3 or 4 contacts per participant. The model fits per round are shown by the median (lines) and 95% interval (shaded), from the first to last participation date of that survey round. The observed activity levels per round (open circles) are placed at the mean participation date of that survey round. Holidays and school holiday periods are shaded in grey.

Next, the model results are extrapolated to the general population over the full course of the study periods (Fig. 4). The saw-tooth pattern is caused by the weekend effect, which is largest for the 0-17 age group. Over time activity levels generally increase in all age groups in both series (Fig. 4). As a comparison, the results of the PiCo study^4^ are plotted, of which rounds 1, 2, 4 and 5 fell within the CoMix study periods. Participants of the PiCo study reported higher activity levels than CoMix participants. The discrepancy is largest for the 0-17 age group, whereas the 65+ age group all but agrees with the PiCo study results.

**Figure 4.**
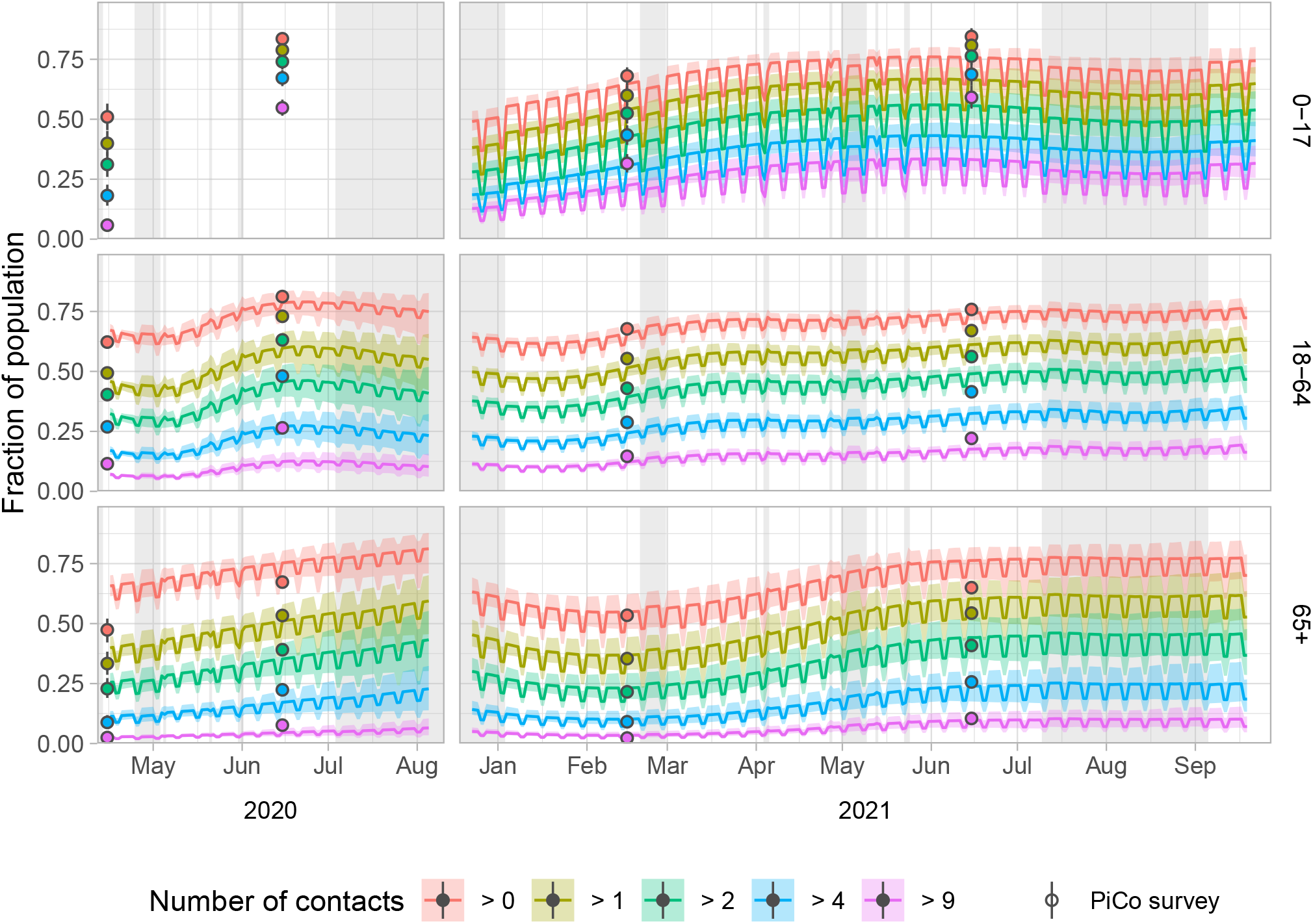
Predicted activity levels over time by series (columns) and age group (rows) for the general population. Activity levels are shown as the fraction of the population that has more than a certain number of non-householdcontacts per day. With five limits (>0, >1, >2, >4 and >9) six activity levels are defined, e.g. the fraction between the limits of >2 and >4 is the activity level that represents 3 or 4 contacts per person. The model predictions are shown by the median (lines) and 95% interval (shaded). The activity levels observed in the contact survey of the independent PiCo study are shown for comparison per round at the mean participation date (points), with bootstrapped 95% confidence intervals. Holidays and school holiday periods are shaded in grey.

## Discussion

In this paper we showed how the number of at-risk contacts per person outside the household increased as the COVID-19 control measures were lifted in the Netherlands, based on the Dutch data of the European CoMix survey.

These findings are consistent with those found in the PiCo studies, even though the activity levels reported in the PiCo data are higher than in the CoMix data. The two studies differed in how the participants report their contacts. In the CoMix study contacts were reported individually with a lot of detail on location, duration, distance from contact, protection measures, etc. Also for the group contacts at work, school, or other places additional details were requested. In the PiCo study a participant reported the total number of contacts in specific age classes, which made reporting many contacts easier. This may partly explain the higher activity levels found in the PiCo study.

We defined six activity levels and used ordinal regression to describe the contact behaviour of the study population. Other studies have used parametric distributions instead, with a maximum number of contacts^22^. This maximum is necessary to yield the same trends in contact behaviour as our results (Supplementary Fig. S3) but also disregards the heavy tails of the distribution of number of contacts. For robustness we also reanalysed the data without distinction between age groups, but using participant age as a covariable over the full age range. Although not markedly different from the main results (Supplementary Fig. S4), this analysis fails to capture any interactions between age group and calendar time.

Participants tend to report fewer contacts when they participate in more rounds. This fatigue effect is also seen in the CoMix data of other countries, but the fatigue effect in the Netherlands is found to be extreme compared to these^29^. A complicating factor in interpreting the fatigue effect is that the participant rounds are not equally distributed over the study period. Most participants have their first participant round in survey round 1 in 2020, and survey rounds 1 and 11 in 2021, due to the study design. In an alternative design used in the UK, the study population is supplemented to reach the target size in every survey round, which leads to a better distribution of the participant rounds over the survey rounds^22^. Also, a lower frequency of the survey rounds may lead to a smaller fatigue effect, but this would conflict with the aim of monitoring contact behaviour close to real time.

To extrapolate our findings to the general population, the fatigue effect needed to be corrected for, and differences between the study population and the general population had to be taken into account. Particularly the age distribution of the study participants varied over the survey rounds, because drop-out rates differed between age groups. Also vaccination and risk status of the participants did not always reflect the vaccination and risk status in the general population. The participants in the age groups of 18-34 years achieved a higher vaccination coverage at an earlier stage than the general population. This could indicate that these groups contained relatively many health care workers, who were vaccinated with priority and may have a higher willingness to get vaccinated. The participants in the age group of 65 years and older lagged behind the general population, because the study population probably did not contain many care home residents who were vaccinated with priority. The fraction of high risk adult participants was higher than in the general population. The risk status was reported by participants themselves and compared to the fraction of the population that is invited for the annual influenza vaccination. Although the high risk definition is the same for both populations, participants may have assessed their risk status differently than their GP would. This is supported by the finding that the high risk participants are mainly overrepresented in the highest age groups in 2020 (Fig. 1B, Tab. 1) when risks may have been perceived to be higher than in 2021 when mass-vaccination against COVID-19 was implemented. This would explain the discrepancy, but it cannot be excluded that they were really high risk participants.

Our findings suggest that the contact data collected in the CoMix survey describe trends in contact behaviour in the general population. As such it can be useful in near real-time monitoring of the transmission potential^22,30^, determining the effect of control measures that are aimed at contact reduction^31,32^, describing determinants for contact behaviour^33,34^, or serving as input for infectious disease models^35,36^. These results can help guide policy in future waves of COVID-19 or other emerging respiratory diseases by monitoring the interaction between contact behaviour, control measures and compliance, and emphasize that contact surveys such as the CoMix study are indispensable for providing a quantitative basis to public health policy.

## Data Availability

All data produced are available online at zenodo.org

http://zenodo.org/record/4790347#.YwTvc3ZByUm/

## Data availability

The datasets analysed in the current study are available in the Zenodo repository (https://doi.org/10.5281/zenodo.4790347)^28^ under CC BY 4.0 license.

## Acknowledgements

The authors would like to thank all participants of the CoMix survey. The CoMix study in the Netherlands was made possible through funding from the European Union’s Horizon 2020 research and innovation programme - project EpiPose (Grant agreement number 101003688). This work reflects only the authors’ view. The European Commission is not responsible for any use that may be made of the information it contains. We are grateful to the EpiPose management team for coordinating the CoMix survey so data collection was made possible in the Netherlands.

## Author contributions statement

P.B, J.E., N.H. and J.W. initiated the study; N.H., P.B., C.J., K.v.Z., J.E. designed the survey questionnaire; A.G., C.J., K.v.Z, P.C, J.W., L.B. collected and cleaned the data; J.B. and L.B. analysed the data; J.B. wrote the manuscript; E.V, C.v.H and D.W. collected and cleaned the PiCo data. All authors reviewed the manuscript and approved the final version for publication.

## References

1. Zhang, J. et al. Changes in contact patterns shape the dynamics of the COVID-19 outbreak in China. Science 368, 1481–1486, DOI: http://doi.org/10.1126/science.abb8001 (2020).

2. Latsuzbaia, A., Herold, M., Bertemes, J. P. & Mossong, J. Evolving social contact patterns during the COVID-19 crisis in Luxembourg. PLoS One 15, e0237128, DOI: http://doi.org/10.1371/journal.pone.0237128 (2020).

3. Quaife, M. et al. The impact of COVID-19 control measures on social contacts and transmission in Kenyan informal settlements. BMC Med 18, 316, DOI: http://doi.org/10.1186/s12916-020-01779-4 (2020).

4. Backer, J. A. et al. Impact of physical distancing measures against COVID-19 on contacts and mixing patterns: repeated cross-sectional surveys, the Netherlands, 2016-17, April 2020 and June 2020. Euro Surveill 26, DOI: http://doi.org/10.2807/1560-7917.ES.2021.26.8.2000994 (2021).

5. Bosetti, P. et al. Lockdown impact on age-specific contact patterns and behaviours, France, April 2020. Euro Surveill 26, DOI: http://doi.org/10.2807/1560-7917.ES.2021.26.48.2001636 (2021).

6. Feehan, D. M. & Mahmud, A. S. Quantifying population contact patterns in the United States during the COVID-19 pandemic. Nat Commun 12, 893, DOI: https://doi.org/10.1038/s41467-021-20990-2 (2021).

7. Tomori, D. V. et al. Individual social contact data and population mobility data as early markers of SARS-CoV-2 transmission dynamics during the first wave in Germany-an analysis based on the COVIMOD study. BMC Med 19, 271, DOI: http://doi.org/10.1186/s12916-021-02139-6 (2021).

8. McCreesh, N. et al. Impact of the Covid-19 epidemic and related social distancing regulations on social contact and SARS-CoV-2 transmission potential in rural South Africa: analysis of repeated cross-sectional surveys. BMC Infect Dis 21, 928, DOI: http://doi.org/10.1186/s12879-021-06604-8 (2021).

9. Drolet, M. et al. Time trends in social contacts before and during the COVID-19 pandemic: the CONNECT study. BMC Public Heal. 22, 1032, DOI: https://doi.org/10.1186/s12889-022-13402-7 (2022).

10. Mossong, J. et al. Social contacts and mixing patterns relevant to the spread of infectious diseases. PLoS Med. 5, e74, DOI: http://doi.org/10.1371/journal.pmed.0050074 (2008).

11. Hoang, T. et al. A Systematic Review of Social Contact Surveys to Inform Transmission Models of Close-contact Infections. Epidemiology 30, 723–736, DOI: http://doi.org/10.1097/EDE.0000000000001047 (2019).

12. Levin, A. T. et al. Assessing the age specificity of infection fatality rates for COVID-19: systematic review, meta-analysis, and public policy implications. Eur J Epidemiol 35, 1123–1138, DOI: http://doi.org/10.1007/s10654-020-00698-1 (2020).

13. Hewitt, J. et al. The effect of frailty on survival in patients with COVID-19 (COPE): a multicentre, European, observational cohort study. Lancet Public Heal. 5, e444–e451, DOI: https://doi.org/10.1016/s2468-2667(20)30146-8 (2020).

14. Blomaard, L. C. et al. Frailty is associated with in-hospital mortality in older hospitalised COVID-19 patients in the Netherlands: the COVID-OLD study. Age Ageing 50, 631–640, DOI: http://doi.org/10.1093/ageing/afab018 (2021).

15. Freedman, V. A., Hu, M. & Kasper, J. D. Changes in Older Adults’ Social Contact During the COVID-19 Pandemic. J Gerontol B Psychol Sci Soc Sci 77, e160–e166, DOI: https://doi.org/10.1093/geronb/gbab166 (2022).

16. Verelst, F. et al. SOCRATES-CoMix: a platform for timely and open-source contact mixing data during and in between COVID-19 surges and interventions in over 20 European countries. BMC Med 19, 254, DOI: http://doi.org/10.1186/s12916-021-02133-y (2021).

17. Vos, E. R. A. et al. Nationwide seroprevalence of SARS-CoV-2 and identification of risk factors in the general population of the Netherlands during the first epidemic wave. J Epidemiol Community Heal. DOI: http://doi.org/10.1136/jech-2020-215678 (2020).

18. Vos, E. R. A. et al. Associations Between Measures of Social Distancing and Severe Acute Respiratory Syndrome Coronavirus 2 Seropositivity: A Nationwide Population-based Study in the Netherlands. Clin Infect Dis 73, 2318–2321, DOI: http://doi.org/10.1093/cid/ciab264 (2021).

19. Hooiveld, M., Heins, M., Hendriksen, J. & Korevaar, J. Aantal mensen met medische indicatie voor vaccinatie tegen COVID-19 (Number of people with medical indication for vaccination against COVID-19). https://www.nivel.nl/sites/default/files/bestanden/1003980.pdf (2021). x[Online; accessed 8 August 2022].

20. RIVM. Cumulatieve opkomst tenminste één COVID-19 vaccinatie naar geboortejaar en week (Cumulative coverage of at least one COVID-19 vaccination per birth year and calendar week). https://www.rivm.nl/covid-19-vaccinatie/cijfers-vaccinatieprogramma (2022). [Online; accessed 14 April 2022].

21. Jarvis, C. I. et al. Quantifying the impact of physical distance measures on the transmission of COVID-19 in the UK. BMC Med 18, 124, DOI: http://doi.org/10.1186/s12916-020-01597-8 (2020).

22. Gimma, A. et al. Changes in social contacts in England during the COVID-19 pandemic between March 2020 and March 2021 as measured by the CoMix survey: A repeated cross-sectional study. PLoS Med 19, e1003907, DOI: https://doi.org/10.1371/journal.pmed.1003907 (2022).

23. Tagore, S. Epidemic Models: Their Spread, Analysis and Invasions in Scale-Free Networks, 1–25 (Springer International Publishing, Cham, 2015).

24. Statistics Netherlands. Bevolking; geslacht, leeftijd en burgerlijke staat, 1 januari 2021. https://opendata.cbs.nl/statline/#/CBS/nl/dataset/7461bev/table (2021). x[Online; accessed 1 July 2021].

25. R Core Team. R: A Language and Environment for Statistical Computing. R Foundation for Statistical Computing, Vienna, Austria (2022).

26. Wood, S. Generalized Additive Models: An Introduction with R (Chapman and Hall/CRC, 2017), 2 edn.

27. RIVM. CoMixNL. https://github.com/rivm-syso/CoMixNL (2022).

28. Backer, J. A., Bogaardt, L. & Wallinga, J. CoMix social contact data (Netherlands). https://zenodo.org/record/7276465#.Y-Sk28nMKUk, DOI: https://doi.org/10.5281/zenodo.7276465 (2022). x[Online; accessed 9 February 2023].

29. Wong, K. L. M. et al. Social contact patterns during the COVID-19 pandemic in 21 European countries – evidence from a two-year study. medRxiv DOI: http://doi.org/10.1101/2022.07.25.22277998 (2022).

30. Jarvis, C. I. et al. The impact of local and national restrictions in response to COVID-19 on social contacts in England: a longitudinal natural experiment. BMC Med 19, 52, DOI: http://doi.org/10.1186/s12916-021-01924-7 (2021).

31. Coletti, P. et al. CoMix: comparing mixing patterns in the Belgian population during and after lockdown. Sci Rep 10, 21885, DOI: https://doi.org/10.1038/s41598-020-78540-7 (2020).

32. Munday, J. D. et al. Estimating the impact of reopening schools on the reproduction number of SARS-CoV-2 in England, using weekly contact survey data. BMC Med 19, 233, DOI: http://doi.org/10.1186/s12916-021-02107-0 (2021).

33. Wambua, J. et al. The influence of risk perceptions on close contact frequency during the SARS-CoV-2 pandemic. Sci Rep 12, 5192, DOI: http://doi.org/10.1038/s41598-022-09037-8 (2022).

34. Wong, K. L. M. et al. Pregnancy during COVID-19: social contact patterns and vaccine coverage of pregnant women from CoMix in 19 European countries. BMC Pregnancy Childbirth 22, 757, DOI: http://doi.org/10.1186/s12884-022-05076-1 (2022).

35. Coletti, P. et al. A data-driven metapopulation model for the Belgian COVID-19 epidemic: assessing the impact of lockdown and exit strategies. BMC Infect Dis 21, 503, DOI: https://doi.org/10.1186/s12879-021-06092-w (2021).

36. Franco, N. et al. Inferring age-specific differences in susceptibility to and infectiousness upon SARS-CoV-2 infection based on Belgian social contact data. PLoS Comput. Biol 18, e1009965, DOI: https://doi.org/10.1371/journal.pcbi.1009965 (2022).

