## Supplementary material for "Dynamics of non-household contacts during the COVID-19 pandemic in 2020 and 2021 in the Netherlands"

### ABSTRACT

The COVID-19 pandemic was in 2020 and 2021 for a large part mitigated by reducing contacts in the general population. To monitor how these contacts changed over the course of the pandemic in the Netherlands, a longitudinal survey was conducted where participants reported on their at-risk contacts every two weeks, as part of the European CoMix survey. The survey included 1659 participants from April to August 2020 and 2514 participants from December 2020 to September 2021.
We categorized the number of unique contacted persons excluding household members, reported per participant per day into six activity levels, defined as 0, 1, 2, 3-4, 5-9 and 10 or more reported contacts. After correcting for age, vaccination status, risk status for severe outcome of infection, and frequency of participation, activity levels increased over time, coinciding with relaxation of COVID-19 control measures.

| year | round | 0-11 | 12-17 | 18-24 | 25-34 | 35-44 | 45-54 | 55-64 | 65+ | total |
|---|---|---|---|---|---|---|---|---|---|---|
| 2020 | target | 0 | 0 | 162 | 235 | 225 | 277 | 249 | 352 | 1500 |
| 2020 | 1 | 0 | 0 | 88 | 227 | 236 | 308 | 277 | 386 | 1522 |
| 2020 | 2 | 0 | 0 | 43 | 176 | 211 | 277 | 255 | 351 | 1313 |
| 2020 | 3 | 0 | 0 | 31 | 145 | 168 | 246 | 237 | 316 | 1143 |
| 2020 | 4 | 0 | 0 | 30 | 134 | 153 | 223 | 212 | 268 | 1020 |
| 2020 | 5 | 0 | 0 | 35 | 131 | 174 | 247 | 231 | 306 | 1124 |
| 2020 | 6 | 0 | 0 | 21 | 137 | 174 | 233 | 228 | 318 | 1111 |
| 2020 | 7 | 0 | 0 | 20 | 103 | 140 | 215 | 195 | 279 | 952 |
| 2020 | 8 | 0 | 0 | 22 | 104 | 142 | 214 | 203 | 284 | 969 |
| 2021 | target | 150 | 150 | 248 | 206 | 163 | 184 | 161 | 238 | 1500 |
| 2021 | 1 | 137 | 145 | 239 | 202 | 158 | 180 | 158 | 232 | 1451 |
| 2021 | 2 | 126 | 126 | 139 | 175 | 136 | 177 | 176 | 257 | 1312 |
| 2021 | 3 | 107 | 115 | 174 | 175 | 136 | 149 | 128 | 187 | 1171 |
| 2021 | 4 | 99 | 104 | 112 | 157 | 110 | 125 | 142 | 213 | 1062 |
| 2021 | 5 | 81 | 95 | 94 | 127 | 89 | 132 | 127 | 197 | 942 |
| 2021 | 6 | 87 | 75 | 110 | 151 | 126 | 118 | 95 | 92 | 854 |
| 2021 | 7 | 73 | 67 | 64 | 126 | 85 | 93 | 115 | 143 | 766 |
| 2021 | 8 | 68 | 76 | 80 | 133 | 102 | 123 | 78 | 103 | 763 |
| 2021 | 9 | 49 | 67 | 78 | 117 | 94 | 86 | 61 | 64 | 616 |
| 2021 | 10 | 40 | 65 | 65 | 104 | 89 | 96 | 71 | 31 | 561 |
| 2021 | 11 | 116 | 154 | 195 | 205 | 160 | 198 | 167 | 260 | 1455 |
| 2021 | 12 | 107 | 133 | 178 | 194 | 155 | 160 | 142 | 208 | 1277 |
| 2021 | 13 | 96 | 115 | 124 | 163 | 121 | 154 | 152 | 233 | 1158 |
| 2021 | 14 | 84 | 98 | 85 | 143 | 117 | 134 | 140 | 228 | 1029 |
| 2021 | 15 | 77 | 86 | 84 | 135 | 117 | 123 | 113 | 188 | 923 |
| 2021 | 16 | 71 | 74 | 50 | 117 | 98 | 117 | 112 | 192 | 831 |
| 2021 | 17 | 59 | 82 | 48 | 115 | 103 | 130 | 119 | 106 | 762 |
| 2021 | 18 | 55 | 69 | 41 | 90 | 65 | 72 | 105 | 181 | 678 |
| 2021 | 19 | 64 | 56 | 43 | 90 | 79 | 73 | 70 | 201 | 676 |
| 2021 | 20 | 47 | 81 | 41 | 86 | 84 | 100 | 87 | 160 | 686 |

**Table S1.** Number of participants per survey round of study series in 2020 and 2021 in eight age groups. The target number of participants was to be achieved in round 1 in 2020, and in rounds 1 and 11 in 2021.

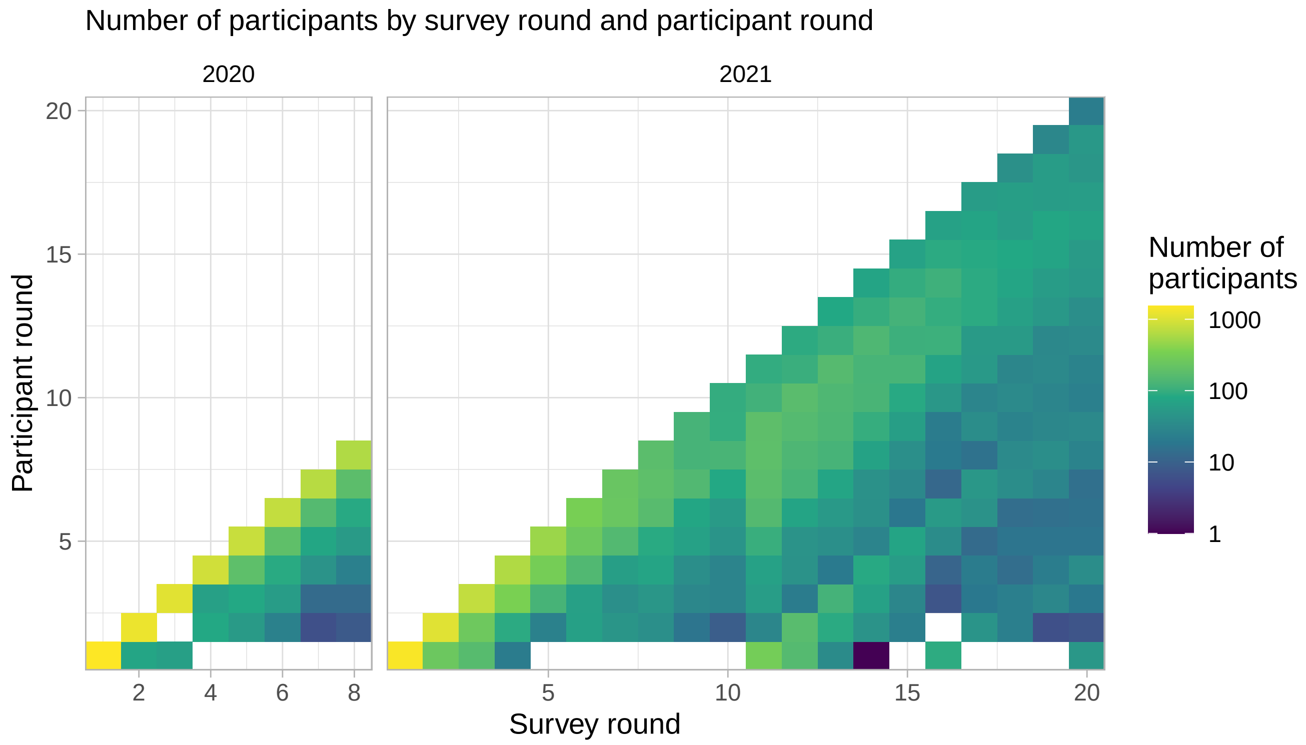


**Figure S1.** Number of participants by survey round and participant round (i.e. the $n^{th}$ time a participant participated). Series 2020 (left) consisted of 8 rounds, series 2021 (right) consisted of 20 rounds. Most participants participated for the first time the start of the series, but some also started during the study period. The panel was supplemented to the target population in round 11 of series 2021.

| series | age_group | Variable | Estimate | Std. Error | t value | p-value | |
|---|---|---|---|---|---|---|---|
| 2020 | 18-64 | (Intercept) | -0.23 | 0.06 | -3.85 | $< 0.001$ | *** |
| 2020 | 18-64 | part_vaccTRUE | 0.00 | 0.00 | | | |
| 2020 | 18-64 | part_elevated_riskTRUE | -0.03 | 0.08 | -0.40 | 0.69 | |
| 2020 | 18-64 | weekendTRUE | -0.21 | 0.06 | -3.67 | $< 0.001$ | *** |
| 2020 | 18-64 | holidayTRUE | 0.07 | 0.10 | 0.72 | 0.47 | |
| 2020 | 18-64 | Deviance explained | 0.16 | | | | |
| 2020 | 65+ | (Intercept) | 0.03 | 0.13 | 0.27 | 0.79 | |
| 2020 | 65+ | part_vaccTRUE | 0.00 | 0.00 | | | |
| 2020 | 65+ | part_elevated_riskTRUE | 0.25 | 0.12 | 2.05 | 0.04 | * |
| 2020 | 65+ | weekendTRUE | -0.32 | 0.08 | -3.89 | $< 0.001$ | *** |
| 2020 | 65+ | holidayTRUE | -0.08 | 0.11 | -0.70 | 0.48 | |
| 2020 | 65+ | Deviance explained | 0.14 | | | | |
| 2021 | 0-17 | (Intercept) | -0.81 | 0.08 | -9.91 | $< 0.001$ | *** |
| 2021 | 0-17 | part_vaccTRUE | 0.12 | 0.19 | 0.64 | 0.52 | |
| 2021 | 0-17 | part_elevated_riskTRUE | -0.15 | 0.27 | -0.56 | 0.57 | |
| 2021 | 0-17 | weekendTRUE | -0.67 | 0.08 | -8.35 | $< 0.001$ | *** |
| 2021 | 0-17 | holidayTRUE | -0.20 | 0.10 | -2.04 | 0.04 | * |
| 2021 | 0-17 | Deviance explained | 0.17 | | | | |
| 2021 | 18-64 | (Intercept) | -0.92 | 0.06 | -16.00 | $< 0.001$ | *** |
| 2021 | 18-64 | part_vaccTRUE | 0.09 | 0.07 | 1.31 | 0.19 | |
| 2021 | 18-64 | part_elevated_riskTRUE | -0.13 | 0.08 | -1.72 | 0.08 | . |
| 2021 | 18-64 | weekendTRUE | -0.27 | 0.05 | -5.88 | $< 0.001$ | *** |
| 2021 | 18-64 | holidayTRUE | 0.06 | 0.06 | 0.94 | 0.35 | |
| 2021 | 18-64 | Deviance explained | 0.19 | | | | |
| 2021 | 65+ | (Intercept) | -0.82 | 0.15 | -5.51 | $< 0.001$ | *** |
| 2021 | 65+ | part_vaccTRUE | 0.08 | 0.19 | 0.40 | 0.69 | |
| 2021 | 65+ | part_elevated_riskTRUE | -0.10 | 0.11 | -0.89 | 0.37 | |
| 2021 | 65+ | weekendTRUE | -0.44 | 0.08 | -5.65 | $< 0.001$ | *** |
| 2021 | 65+ | holidayTRUE | 0.04 | 0.10 | 0.42 | 0.68 | |
| 2021 | 65+ | Deviance explained | 0.16 | | | | |

**Table S2.** Results for fixed effects of generalised additive model, by series (2020, 2021) and age group (0-17, 16-64, 65+). Included fixed effects are participant vaccination status (part_vacc), participant risk status (part_elevated_risk), weekend and holiday. Holidays include general holidays and school holidays. The last row of each section refers to the explained deviance. Significance levels: *** ($p$-value $< 0.001$), ** ($p$-value $< 0.01$), * ($p$-value $< 0.05$), . ($p$-value $< 0.1$)

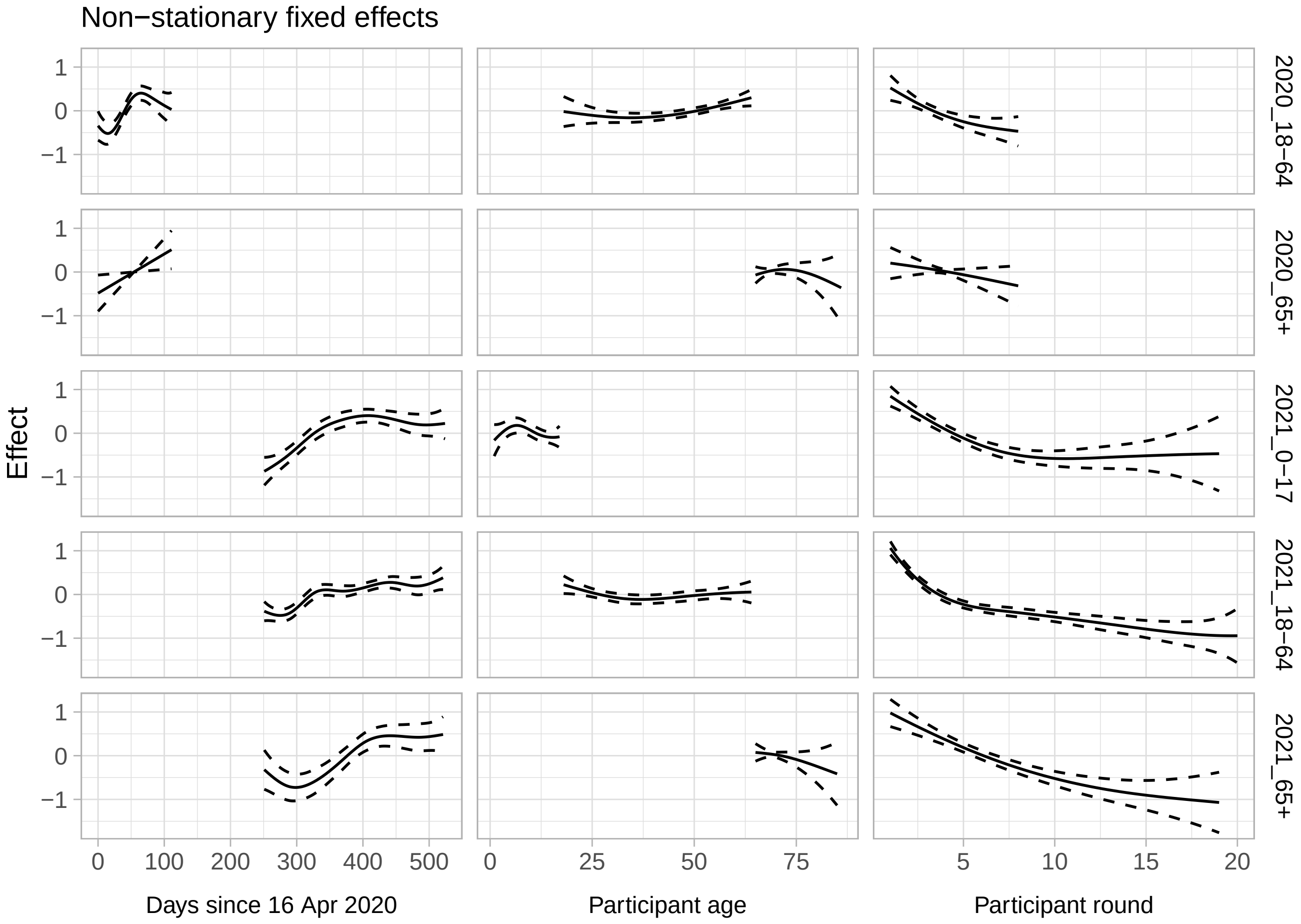


**Figure S2.** Results for non-stationary fixed effects of generalised additive model, by series (2020, 2021) and age group (0-17, 16-64, 65+). Calendar time (expressed as days since first survey), participant age, and participant round (i.e. the $n^{th}$ time a participant participated) are included as cubic splines. Shown are the fitted splines (solid lines) $\pm$ the standard error (dashed lines).

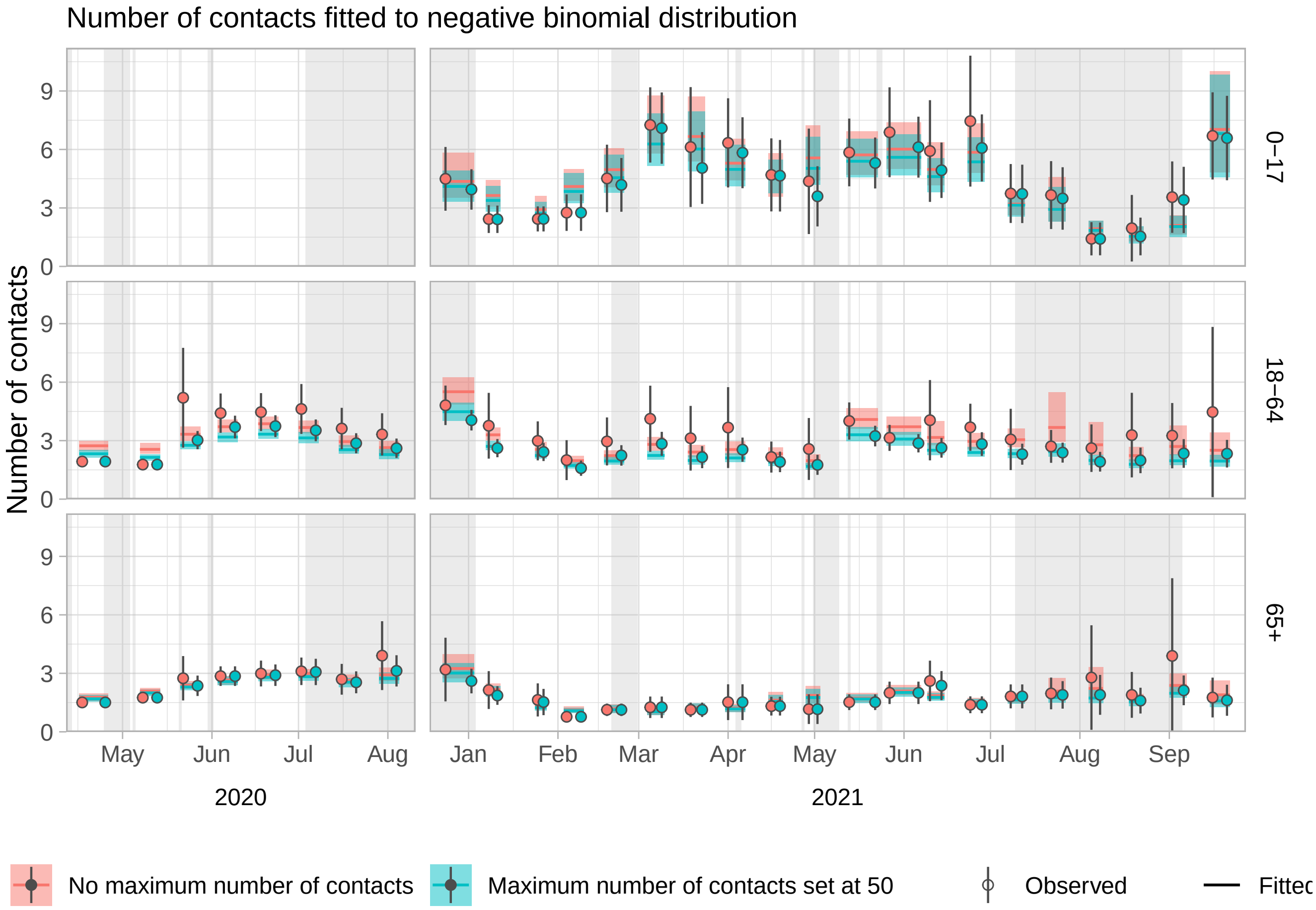


**Figure S3.** Fitted and observed number of contacts over time by series (columns) and age group (rows), with and without setting a maximum for the number of contacts. The model fits per round are shown by the median (lines) and 95% interval (shaded), from the first to last participation date of the survey round. The mean observed number of contacts, without and with set maximum are placed at the start and end of the survey round (circles, with 95% CI errorbars). Holidays and school holiday periods are shaded in grey. Only 0.8% of the observations are higher than 50, but they lead to much higher and more uncertain estimates. Setting a maximum yields the same trends in contact behaviour as our results but also disregards the heavy tails of the distribution of number of contacts.

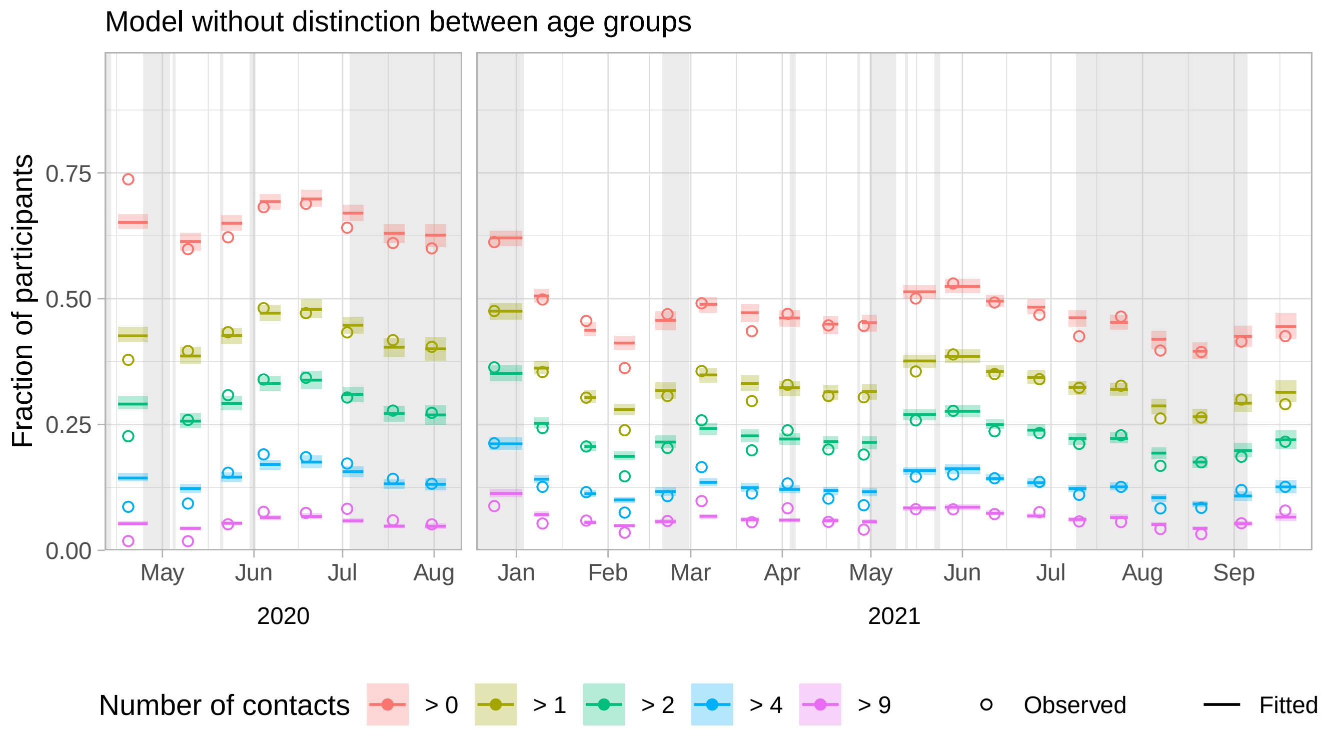


**Figure S4.** Fitted and observed activity levels over time by series. Activity levels are shown as the fraction of participants that report more than a certain number of non-household contacts per day. With five limits (>0, >1, >2, >4 and >9) six activity levels are defined, e.g. the fraction between the limits of >2 and >4 is the activity level that represents 3 or 4 contacts per participant. The model fits per round are shown by the median (lines) and 95% interval (shaded), from the first to last participation date of that survey round. The observed activity levels per round (open circles) are placed at the mean participation date of that survey round. Holidays and school holiday periods are shaded in grey. The model presented in the main text distinguishes between 3 age groups (0-17, 18-64, and 65+), while here they are analysed together. Participant age is modelled with a spline over the full age range. Although not markedly different from the main results, this analysis fails to capture any interactions between age group and calendar time, for instance when the oldest age group increased their contacts in 2021 later than the rest of the population (see Fig. S2).
